# An analysis of international strategies for monitoring and preventability described in medicinal product information: a study protocol

**DOI:** 10.1101/2024.06.14.24308939

**Authors:** Daniele Sartori, Jeffrey K. Aronson, G. Niklas Norén, Igho J. Onakpoya

## Abstract

**Introduction:** Product information is intended to be a reference for healthcare professionals to verify instructions for monitoring and preventability of adverse drug reactions (ADRs), among other things. International comparisons of these documents, using the Systematic Information for Monitoring (SIM) method, have highlighted discrepancies in the instructions for monitoring, but there has been no comparison of preventability instructions.

**Objectives:** To quantify and compare, across different countries, the completeness of instructions for monitoring and preventability provided to healthcare professionals in medicinal product information.

**Methods:** We shall retrieve information included with medicinal products that have been involved in signals communicated by regulators, in 2014–2019 and based on clinical assessments of reports of ADRs, from the websites of 35 regulatory agencies. We shall evaluate the completeness of instructions for monitoring using a modified version of the SIM method; a score of 67% will qualify a monitoring instruction as sufficiently complete. To evaluate the completeness of instructions for preventability, we have derived a framework from the Dose-responsiveness-Temporality-Susceptibility (DoTS) classification of ADRs and related implications, comprising domains and items/implications. We shall iteratively develop a threshold to define the sufficiency of completeness of instructions based on data distribution across DoTS domains. We shall present descriptive statistics by country for each item of the framework and by total scores, using tables, or figures where necessary.

**Outcomes:** Our target audience is regulators, and the results should highlight gaps in the level of information available to healthcare professionals. This study may also provide some insights into how suspicions of causality that arise from clinical assessments of reports of ADRs translate into actionable recommendations in clinical practice.

## Introduction

International comparisons of information provided with different medicinal products concerning the same active ingredient showed discrepancies in categorization, types and counts of adverse drug reactions (ADRs) across harmonized sections of the documents (e.g. warnings or boxed warnings, precautions, contraindications, or untoward effects) [1-6].

Structured analyses of the instructions for monitoring of ADRs have also highlighted cross-country differences and lack of completeness for, or actionability in, clinical practice [7, 8]. Appealing to differences in regulatory practices may only superficially explain these findings, while further enquiries into the strength of evidence that underpins decision-making may better contextualize them and highlight their public health impact [9].

Amendments to the recommendations in product information may be required, based on assessments of evidence that suggest causal relationships between medicinal products and ADRs, that is, signals of ADRs [10]. A recent scoping review of studies of signals showed that clinical assessments of reports of ADRs supported over 2000 signals in the past 40 years [11], consolidating the role of this type of evidence in pharmacovigilance. Despite their volume there are limited insights into how signals translate into the recommendations available in product information [12-14].

While analyses of recommendations for monitoring are available [7, 8], not all ADRs can be monitored: strategies for monitoring may not be feasible (continuously or at intervals), cost-effective, or have sufficient sensitivity or specificity [15]. Preventive strategies such as avoidance of treatment in at-risk populations, dose reduction, and treatment withdrawal, may be recommended when monitoring is unfeasible, but their availability, actionability, and completeness remain to be evaluated.

This study therefore aims at investigating the extent of international discrepancies in regulation, and its objectives are to 1) quantify the completeness of the instructions for monitoring and preventability of ADRs in product information intended for healthcare professionals, and 2) compare it across regulatory agencies or authorities.

## Methods

### Initial basis of the study data

Using previously published datasets of signals of ADRs [11, 16], we shall retrieve, for each signal: (a) the medicinal products (active ingredient, coded to WHODrug Global [17], and defined by [18]) and their earliest launch dates; (b) adverse events involved (coded to MedDRA, the Medical Dictionary for Regulatory Activities®); (c) the country of communication. To minimize the effect of strength of evidence on the known discrepancies of product information [9], we shall restrict the evidence-base of signals to those categorized as arising solely from clinical assessments of reports of ADRs. We shall also require signals to have been communicated by regulatory agencies or authorities, or by national and regional pharmacovigilance centers in 2014–2019 to allow for at least 5 years to have lapsed before any amendments to product information could be made.

### Data retrieval

We consider product information as documents produced by regulatory agencies, intended for healthcare professionals, that describe the properties of medicinal products. Some such documents include Summaries of Products Characteristics (SmPCs) in the European Union; this term is synonymous with Labeling/Package Inserts in the United States of America and Data Sheets in New Zealand.

We shall use the earliest launch date of each medicinal product to determine their status as reference products and obtain the most recent version (as of 06/2024) of their product information in English from the websites of 34 regulatory agencies or authorities, listed in Appendix 1. The list has been adapted from previous systematic reviews [11, 14]. We do not place restrictions on the marketing status of a medicinal product, so withdrawn medicinal products are included as they may still be available on some markets worldwide [14]. Our focus on reference products is driven by observations of discrepancies between their product information and that of their generic or biosimilar counterparts [19, 20], and on the assumption that a longer time on market would result in more complete documents. Where product information is available only for generic or biosimilar formulations, we shall retrieve data on the most recently updated version, as reported on the available date of update.

Product information of medicines marketed as part of national procedures in the European Union may not be available from the website of the European Medicines Agency and will be obtained from the United Kingdom (UK) Medicines and Healthcare products Regulatory Agency (MHRA), the Republic of Ireland (IE) Health Products Regulatory Authority (HPRA). For products licensed outside the UK or IE, we shall use the websites of the Italian Medicines Agency or the Malta Medicines Authority. For signals of ADRs reported at a pharmaceutical class level, we shall consider the product information of the reference medicinal product with the earliest launch date, or the most up-to-date version of the documents on the relevant generic or biosimilar.

### Data extraction

#### Overarching considerations for data extraction

To identify terms and textual sections of the product information that best concern the ADRs involved in the signals, we shall make allowance for broad clinical classifications that may subsume some symptomatic ADRs [21]. For example, the expression “hypersensitivity reactions” may subsume ADRs of “anaphylaxis”, “allergic reaction”, or “angioedema”; thus, instructions for monitoring relevant to hypersensitivity will apply, by extension, to signals of allergic reactions. This approach will also guide the extraction of ADR terms that are not identical to those reported in the communicated signals but may otherwise be deemed as documented in the product information.

For each ADR of the included signals, we shall search the product information for: (a) information on their clinical aspects; (b) instructions for monitoring and (c) preventability. We shall enter the data into a customized Excel spreadsheet, grouped by type of instruction, category of clinical aspect, signal of ADR, section of the product information (e.g. warnings, precautions etc.) and country of origin. We describe, in the sections below, the frameworks that will guide data extraction and categorization of relevant clinical aspects and instructions for monitoring or preventability.

#### Establishing a framework for guiding the extraction of clinical aspects of the ADRs and of instructions for in-principle preventability

Within the scope of preventability, we include precautionary, preventive, and mitigating actions [22] and consider preventability in the absence of errors, i.e. in-principle preventability. The preventability of ADRs is typically assessed against criteria derived empirically or from principles [23]. One method (“EIDOS” and “DoTS” mnemonics) combines the evaluation of mechanistic and clinical aspects of ADRs to propose strategies for preventability [15]. It notes that an *E*xtrinsic species (a medicine) may interact with an *I*ntrinsic species (e.g. a receptor) when they are *D*istributed together (i.e. in the same milieu), to produce an *O*utcome (the adverse effect) with its *S*equelae (the adverse reaction). Furthermore, it accounts for the *Do*se-relatedness of outcomes (collateral, toxic, or hypersusceptibility effects), their *T*ime course (e.g. first dose), and their *S*usceptibility factors (e.g. genetic, age, sex, etc.) [15, 23-25]. DoTS pairs these three components to dependent categories of ADRs, and to the categories assigns implications for preventability. EIDOS/DoTS have been used for devising preventive studies in pharmacovigilance planning [26], to verify the justifiability of withdrawals [27], and to evaluate the in-hospital preventability of ADRs [28, 29].

We repurpose the three categories of DoTS as domains of interest for classifying the information on the clinical aspects of the ADRs, adopt the dependent categories as items of interest to guide the extraction of the clinical aspects of the ADRs that appear in product information, and the related implications to guide instructions for preventability. Beyond extrinsic species and sequelae, clinical assessments of reports of suspected ADRs rarely provide sufficient evidence on the pathophysiological mechanism of an ADR, thus we retain only the DoTS component of the method. In Table 1, we report the framework we shall use for data extraction.

**Table 1.**
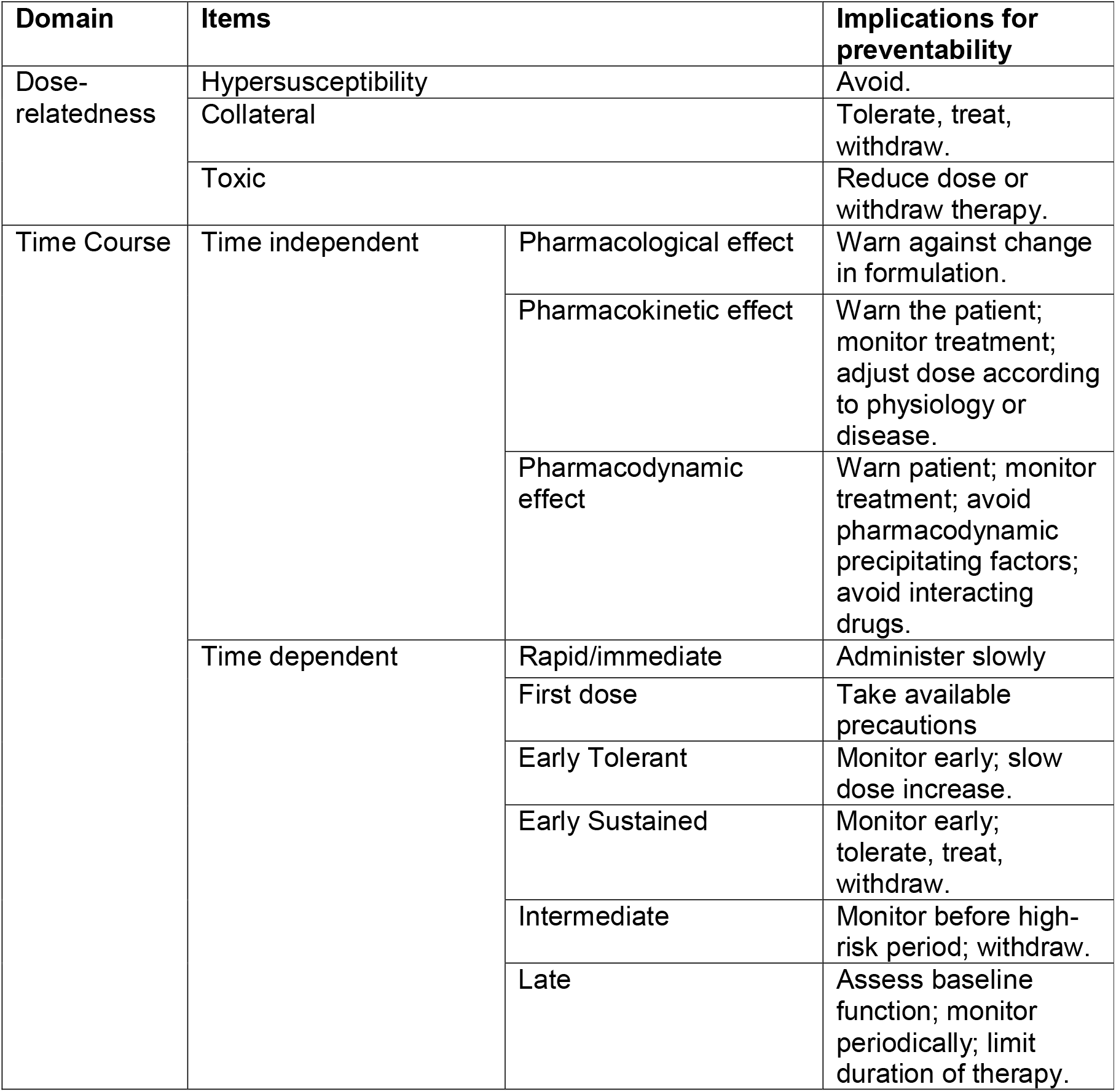
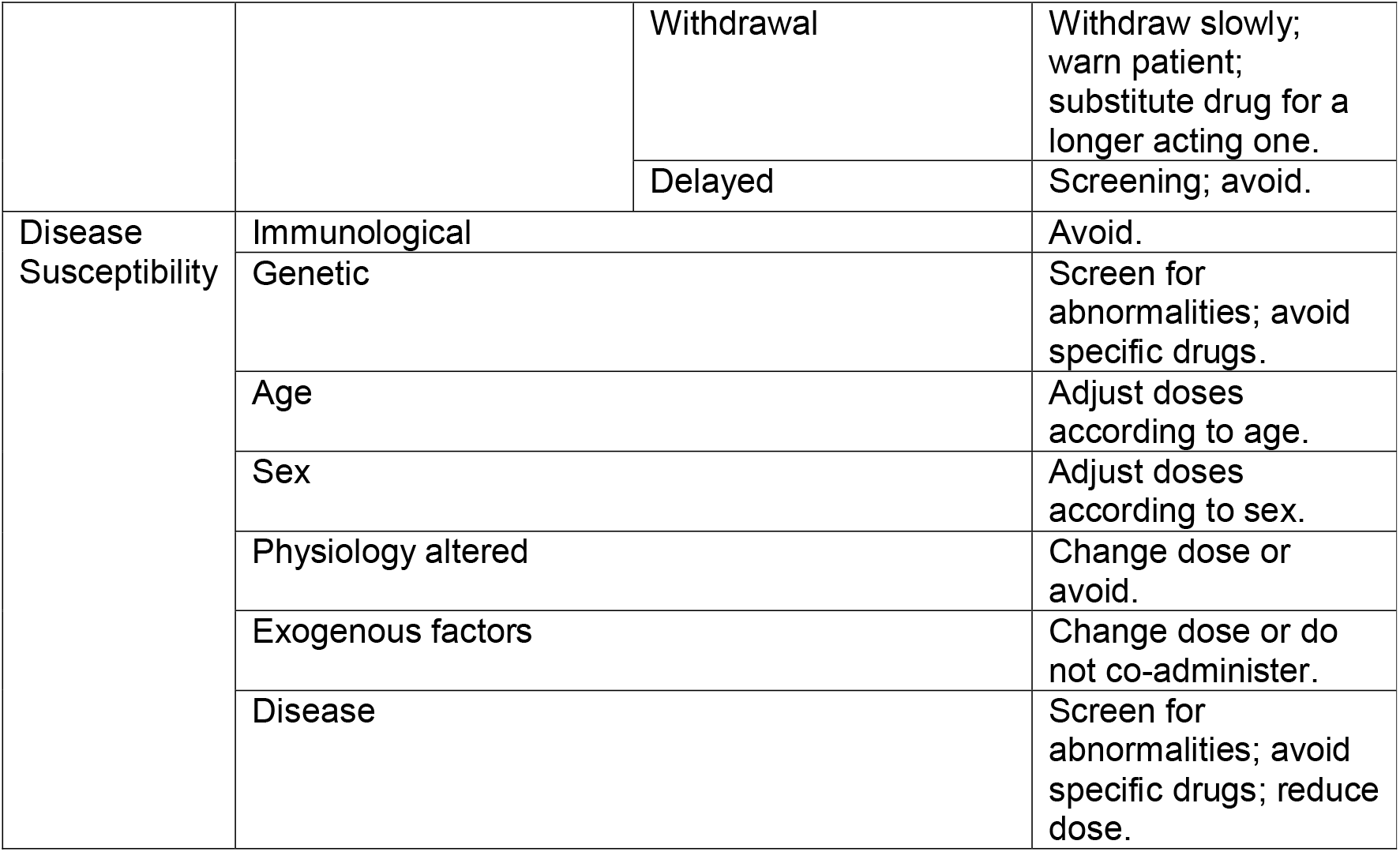
Domains of the DoTS classification, with corresponding items and examples of implications for preventability, derived from [4, 16, 21, 22]. Items will guide the extraction of clinical features of the ADRs from product information, and the implications of instructions for preventability, while the Domains will guide their classification.

#### Systematic Instructions for Monitoring: extracting information relating to instructions for monitoring

Monitoring is the “repeated testing aimed at guiding and adjusting the management of a chronic or recurrent condition” [30]. We consider the monitoring of acute or chronic ADRs at the patient-level as a ‘planned and systematic assessment of adverse effects’ [31]. Both point-of-care (e.g. signs and symptoms of a condition, heart rate, etc.) and laboratory-based monitoring (e.g. white blood cell counts) in different settings (i.e. primary, intensive, and ambulatory care), fall under the scope of this research. Surveillance of healthcare products (qualified with “passive”, “active” and “stimulated”), or population-level monitoring, is distinct from monitoring and is not part of this research [32].

We shall rely on a modified version of the Systematic Instructions for Monitoring (SIM) method to guide the extraction of this type of instructions from product information [7, 8]. In its original conception, the method includes seven guiding questions, whereas we adopt six. As shown in Table 2, we adopt a Domain/Items structure, like that of our proposed framework above, to classify instructions for monitoring (the items) under guiding questions (the domains). The modified SIM method has been applied to highlight the deficiencies of product information or Direct Healthcare Professional Communications in reporting recommendations for the monitoring of ADRs [7, 8, 33].

**Table 2.** Domains of the SIM method, together with examples of items that can be deemed as complete to formulate instructions for monitoring, derived from [8].

| Domain | Examples of items |
| --- | --- |
| What to monitor | Heart rate |
| When to start monitoring | Monitor heart rate before start of treatment |
| When to stop monitoring | Stop monitoring one week after initiation |
| How frequently to monitor | Every 30 days |
| What to look for/Critical value | Heart rate > 100 |
| How to respond | If heart rate > 100, discontinue treatment immediately |

### Data analysis

#### Summary descriptive statistics

For each signal, we shall record the number of product information that do and do not record the relevant ADRs, together with the totals of product information that contain instructions for monitoring and preventability. We shall present these findings in tables grouped by country of origin of the product information, and/or by type of medicinal product.

#### Assessment of completeness of the instruction for monitoring and in-principle preventability

Unlike the 35-point system of the original, the modified SIM method entails scoring any extracted information as either complete or incomplete, awarding a complete item, and its corresponding domain, 1 point. A total score of 4 (67%) across all 6 domains identifies a product information as having “sufficiently complete” monitoring instructions for a given signal of ADR [8].

To design a scoring system for assessing the completeness of in-principle preventability instructions we shall initially assign data to three categories: available item; available instruction(s); co-occurrence of item and instruction(s) matching the implication of the corresponding DoTS domain. Based on the data distribution, we shall iteratively derive a score that accounts for the possible texts’ granularity and determine a threshold above which one may consider preventability instructions as sufficiently complete.

Because scoring systems entail subjectivity, one researcher (DS) will score the data, and another (IJO) cross-validate the findings. Disagreements will be resolved through discussions and a third (JKA) shall arbitrate if no consensus can be reached.

We shall present SIM scores, and scoring of preventability instructions, using medians and interquartile ranges in tabular formats or plots, as appropriate, grouped by country of origin of product information.

#### Cross-country comparison

After assigning scores we shall compare the completeness and types of preventability and monitoring instructions available in product information across countries, through descriptive statistics, e.g. frequencies and medians/interquartile ranges, presented in tables or plots (UpSet, or bar and whisker plots), where necessary.

Before prescribing a specific statistical test to estimate quantitative difference in scores across countries, we shall analyse the distributional properties of the scores for variance and evaluate normality and the risk of multiple comparisons. We shall therefore visualize the data on scatter plots and subsequently determine a fitting statistical test, sample size permitting.

## Discussion/Conclusions

This study is intended to provide regulators with an international overview of the state of the quality of recommendations in product information, as well as indications that may merit clearer formulations to benefit healthcare professionals. It may also inform readers about the nature of regulatory actions that may follow signals based on clinical assessments of reports of ADRs. We plan to disseminate the findings of this study as a peer-reviewed publication and as part of Daniele Sartori’s DPhil thesis with the University of Oxford. We may present part of the results as part of conference proceedings, abstracts, or posters.

We acknowledge that our choice of interval in the years of communication (2014 – 2019) may have allowed for additional evidence to have accrued before any instructions were added to the product information. As such we may be unable to show a direct causal link between the communication of a signal and addition of recommendations to the product information.

We have prespecified a list of regulatory agencies from which to obtain product information, but some not be publicly available. In addition, we shall only consider product information in English. These choices may result in the omission of product information that would otherwise contain recommendations eligible for evaluation. Our list should be sufficiently wide to ensure acceptable coverage across world regions, as it has before [11, 14, 27].

We further recognize that not all ADRs may be accompanied by instructions for monitoring or preventability; for example, clinically mild ADRs without evidence of effect on quality of life may be merely listed as possibly occurring, as could those for which the evidence upon communication was insufficient to draft any recommendations at all. We plan to address this possible limitation of the study in our discussion, contextualizing our findings with notes on feasibility of monitoring or preventive strategies and with the guidelines for compiling product information published by regulatory agencies [34, 35].

## Data Availability

We will abide by data availability requirements of the journal to which we will submit the study this protocol describes. If there are no specific requirements, all data produced by the study described in the protocol will be made available upon reasonable request to the authors.

## Disclosures

All authors have completed the International Committee of Medical Journal Editor forms. This work is part of DS’s DPhil in Evidence-Based Health Care at the University of Oxford, funded by the Uppsala Monitoring Centre (DS’s employer). No undue influence or coercion was exerted on DS by the funder throughout the design and conception of the study, or the writing of this protocol.

DS, IJO and NN have no relevant conflicts of interest. JKA has published papers and edited textbooks on adverse drug reactions and has published papers dealing with the definitions of relevant terms.

MedDRA® the Medical Dictionary for Regulatory Activities terminology is the international medical terminology developed under the auspices of the International Council for Harmonisation of Technical Requirements for Pharmaceuticals for Human Use (ICH).

MedDRA® trademark is registered by ICH. This study protocol does not reflect the opinions of the World Health Organization.

## Ethics Approval

This research does not involve human participants, tissues and/or personal data. As such, ethics approval was not required.

### Appendix 1

#### List of Regulatory Agencies or Authorities from which product information or equivalent documents will be retrieved

1. Australia: Therapeutic Goods Administration (TGA).
2. Bahamas: Bahamas National Drug Agency (BNDA).
3. Bhutan: Drug Regulatory Authority.
4. Botswana: Ministry of Health, Republic of Botswana.
5. Canada: Health Canada Drug Product Database (DPD).
6. European Union: European Medicines Agency (EMA).
7. Gambia: Medicine Regulatory Authority.
8. Ghana: Ghana Food and Drugs Authority.
9. Hong Kong: Hong-Kong’s Department of Health Drug Office.
10. India: Central Drugs Standard Control Organization (CDSCO).
11. Jamaica: Ministry of Health, Kingston, Jamaica.
12. Japan’s Pharmaceuticals and Medical Devices Authority.
13. Kenya: Pharmacy and Poisons Board, Kenya.
14. Korea (Republic of): Korea Institute of Drug Safety and Risk Management.
15. Liberia: Liberian Medicines Health Products Regulatory Authority.
16. Malaysia: National Pharmaceutical Regulatory Agency.
17. Namibia: Namibia Medicines Regulatory Council.
18. New Zealand: New Zealand Medicines and Medical Devices Safety Authority.
19. Nigeria: National Agency for Food and Drug Administration and Control (NAFDAC).
20. Pakistan: Drug Regulatory Authority of Pakistan (DRAP).
21. Philippines: Food and Drug Administration, Philippines.
22. Rwanda: Food and Drug Authority.
23. Saudi Arabia: Saudi Food and Drug Administration.
24. Sierra Leone: Pharmacy Board of Sierra Leone.
25. Singapore: Health Sciences Authority.
26. South Africa: South Africa Medicines Control Council.
27. Sri Lanka: Cosmetics, Devices and Drug Regulatory Authority.
28. Tanzania: Tanzania Medicines & Medical Devices Authority.
29. Thailand: Food and Drug Administration, Thailand.
30. Trinidad and Tobago: Ministry of Health, Trinidad and Tobago.
31. Uganda: National Drug Authority.
32. United States of America: Food and Drug Administration (FDA).
33. Zambia: Pharmaceutical Regulatory Authority (PRA).
34. Zimbabwe: Medicines Control Authority of Zimbabwe (MCAZ).

